# EEG coherence as a marker of Alzheimer’s disease

**DOI:** 10.1101/2022.07.24.22277966

**Authors:** Dina Radinskaia, Crystal Radinski

## Abstract

**Background:** Progressive deterioration of synaptic plasticity and synaptic connectivity between neurons is a neurophysiological hallmark of brain ageing and has been linked to the severity of dementia. We hypothesized that if synaptic disconnection as the neuropathology of Alzheimer’s disease (AD) is responsible for the failure of the brain to integrate various regions into effective networks, then electroencephalographic evidence of the disruption of functional connectivity might be used to diagnose Alzheimer’s dementia. We proposed that changes in EEG coherence, a measure of functional interaction between the brain collaborating areas, can be detected in a clinical setting and serve as a marker of neuronal disconnection. Improving the accuracy and reducing the time needed to diagnose AD could allow timely interventions, treatments, and care cost reduction.

**Methods:** This study examined group differences in EEG coherence within global cortical networks at rest and during executive challenges among patients with AD, individuals with mild cognitive impairment, and healthy controls.

**Results:** Decreased EEG coherence has been discovered in cross-hemisphere frontal, temporal, parietal and occipital pairs in the AD group at rest and when challenged with tasks requiring comprehension, analysis, perceptual-motor response, and executive functioning. The most notable changes were detected in F3-F4 Beta with the visual-spatial task challenge, P7-P8 Beta during the writing task, T7-T8 Gamma during a task requiring speech understanding and O1-O2 Alpha during orientation in space task.

**Conclusions:** The study identified several potential EEG biomarkers of AD. More research is needed to identify sensitivity and specificity of the markers.

## 1. Introduction

Alzheimer’s disease (AD) is the most common form of major neurocognitive disorder in older adults. In the United States, AD is the sixth leading cause of death, killing more people than breast cancer and prostate cancer combined [1]. Clinicians need to be able to accurately diagnose and manage the early cognitive manifestations of AD, particularly as new therapies are developed.

A definite diagnosis of AD can be established only in the presence of histopathologic evidence [2]. As a probable diagnosis, AD is evaluated by a series of clinical and neurophysiological examinations repeated over a period of time and demonstrating progressive cognitive decline present in at least one area of cognitive domains. Patients and families are often uncertain about the onset of symptoms since the initial manifestations of dementia are discrete and inaccurately ascribed to “ageing.” Identifying AD is a time-consuming process, and diagnosis is often missed. One study found that the diagnosis was missed in 21% of demented or delirious patients on a general medical ward, while 20% of non-demented patients were mistakenly diagnosed [3].

Although the neuropathology of AD (neurofibrillary tangles, amyloid plaques, and synaptic dysfunction) has been closely studied, the pathophysiological foundation of cognitive impairment is less clear [4]. Executive function is very complex and relies on the coordination of multiple brain regions. Synaptic dysfunctions were detected in the early stages of dementia even before the emergence of any symptoms [5, 6]. It has been hypothesized that the disconnection between regions due to the brain’s synaptic dysfunctions could disrupt functional connectivity and result in the brain’s failure to integrate various regions into effective networks [7-9]. Progressive deterioration of synaptic plasticity and synaptic connectivity between neurons is a neurophysiological hallmark of brain ageing and has been linked to the severity of dementia [10]. Compensatory remodelling ensures functional maintenance of neurons and constitutes brain reserve. Therefore, neurodegeneration may occur in the absence of symptoms for an uncertain period of time. The onset of functional deterioration in AD is often uncertain, as many diseases could cause transient functional decline. The use of EEG markers of AD in conjunction with standard assessments of cognitive functions with neuropsychological batteries could help detect neuronal dysfunction and decreasing brain reserve and thus facilitate earlier recognition of brain neurocognitive disorder.

Numerous studies have examined functional connectivity in AD with EEG [11-13]. EEG coherence represents the functional interaction between two regions [14, 15]. It is an advantageous method for exploring neuronal network functioning and could help test the disconnection hypothesis. In studies examining resting synchronization in AD patients, a global reduction in alpha and beta coherence was reported as the most common finding [6, 9, 13]. Decreased coherence has been discovered in cross-hemisphere frontal, temporal, parietal and occipital pairs [12, 16-18], as well as decreased long-distance frontoparietal intrahemispheric coherence [10, 19, 20].

Several studies have examined task-related EEG coherence in patients with AD and MCI (Mild Cognitive Impairment). Task-related coherence is the measure of coherence during the performance of a cognitive task and is reflective of the sensory and cognitive networks activity [7, 11, 15]. In AD patients, EEG coherence has been examined during the performance of visual tasks [21, 22], short-term memory tasks [23, 24] and target counting [25]. Decreased EEG coherence was more extensive in AD patients during task performance than during control (rest) [21, 25]. During a visual task, fronto-posterior coherence was reduced in AD patients in the delta, theta and alpha bands [21, 22]. During short-term memory tasks, coherence was decreased in the alpha and beta bands [23, 24]. The decrease in both alpha and beta band task-related EEG coherence in the frontal and temporal interhemispheric regions in patients with AD was demonstrated in multiple studies and seemed to be a potential marker [15, 22, 24, 26]. We hypothesized that if synaptic disconnection as the neuropathology of AD is responsible for the failure of the brain to integrate various regions into effective networks, then electroencephalographic evidence of the disruption of functional connectivity might be used to diagnose Alzheimer’s dementia. We explored the relationship between EEG coherence and executive function in patients with MCI, AD and healthy controls. The EEG data were collected during brief cognitive testing comparable to standard screening done in primary care, such as the Mini-Mental State Examination (MMSE) and Montreal Cognitive Assessment Scale (MoCA). We predicted a decrease in alpha and beta band task-related EEG coherence in the frontal, parietal and temporal regions. The study’s general goal was to identify potential AD markers that could facilitate early recognition of neural functional deficiency and reduce the time needed to diagnose major neurocognitive disorders in primary care.

## 2. Methods

### 2.1 Participants

The study evaluated three groups of ten participants with different conditions of cognitive function levels: individuals with normal cognitive function (control), individuals diagnosed with AD and individuals with MCI. Participants were recruited from the community care centers and long-term care facilities in Calgary, Alberta. All participants were between the age of 65 and 85, had at least a grade eight education and were fluent in English. The neurocognitive status of all participants was confirmed within three months before the study by the Memory Clinic team in Calgary through a series of functional and cognitive testing repeated at least three months apart. AD group had the clinical diagnosis also supported by brain MRI results. Participants with unstable medical conditions that might affect cognition (e.g. uncontrolled thyroid dysfunction, B12 deficiency, alcohol abuse) or current (within two weeks) psychotropic medication (e.g. anticholinergics, neuroleptics and benzodiazepines) use were excluded. Participants with stable chronic conditions were recruited for the study. Out of 30 participants, there were two members with a history of NSTEMI, eight with controlled hypertension, six with controlled hypothyroidism, twelve with osteoarthritis, and five with GERD. All participants provided written informed consent. Participants’ age, gender, and educational level did not significantly differ across the three groups. Global Deterioration Scale, Mini-Mental State Examination, and Montreal Cognitive Assessment Scale were used to document all participants’ cognitive status (Table 1).

**Table 1.** Descriptive information of study groups.

| Characteristics | Control<br>(n=10) | AD<br>(n=10) | MCI<br>(n=10) | Control vs AD<br>P-value |
| --- | --- | --- | --- | --- |
| Age, years (mean±SD) | 78.4±5.08 | 79.7±4.32 | 80.1±1.44 | 0.27 |
| Education, years (mean±SD) | 12.2±2.2 | 13.4±2.31 | 11.8±1.75 | 0.12 |
| Female gender (n, %) | 6 (60) | 6 (60) | 5 (50) | 0.50 |
| GDS (1-7) | 1±0 | 4±0 | 2.1±0.31 |  |
| MMSE (1-30) | 28.6±0.67 | 20.6±0.51 | 23±0.31 |  |
| MoCA (1-30) | 27±0.87 | 16.9±0.73 | 22±0.66 |  |

AD, Alzheimer's disease; MCI, mild cognitive impairment; GDS, Global Deterioration Scale; MMSE, Mini-Mental State Examination; MoCA, Montreal Cognitive Assessment Scale.

The Research Ethics Office of the University of Alberta, Canada, reviewed and approved this study’s adherence to ethical guidelines (HREBA.CHC-16-0053).

### 2.2 Cognitive state tasks panel

Components from the MoCA and MMSE, commonly used clinical screening tests, were used to create a Cognitive State Tasks Panel (CSTP) for the study examining those cognitive domains most affected in AD: orientation to time and place, registration, attention and calculation, repetition, complex commands, recall and language (Table 2).

**Table 2.** Cognitive State Tasks Panel

| # | Question | Test/syndrome | localization |
| --- | --- | --- | --- |
| 1 | What is the year now? Season? Month? Date? Day? | orientation | altered mental state/<br>cortex global |
| 2 | Where are we now:<br>What country?<br>Province? City? Place? Floor? | orientation | altered mental state/<br>cortex global |
| 3 | I will give you three words to memorize;<br>Please repeat them after me and I will ask<br>you about them later again<br>APPLE PENNY TABLE | Short term memory | the pre-frontal lobe |
| 4 | Could you please count backward from<br>100 by 7: 93""86""79""72""65""58""51 | Acalculia | the left angular gyrus<br>and<br>the frontal lobe |
| 5 | Earlier I gave you three words to<br>memorize;<br>Could you recall them please? | Long term memory | the frontal cortex |
| 6 | Please name objects I point at:<br>PEN WATCH | Anomia<br>aphasia | Broca's area<br>( frontal lobe) |
| 7 | Please repeat after me<br>"THE SKY IS BLUE" | Broca's aphasia | left inferior frontal |
| 8 | Please listen carefully and do as I say:<br>"Take the paper in your right hand, fold it<br>and place it on the table" | Wernicke's aphasia<br>Apraxia | left superior temporal<br>and inferior parietal<br>region |
| 9 | Please read this and do what it says (written instructions) | comprehension/Wernicke's aphasia | left superior temporal and inferior parietal region |
| 10 | Please write a sentence about anything | Agraphia<br>apraxia | the left superior frontal and parietal areas |
| 11 | Please copy the picture | Visuospatial/<br>Constructional apraxia<br>/Ideational apraxia | the medial temporal and parietal lobes |
| 12 | Please follow the pattern | Executive function/<br>Ideational apraxia | the medial temporal and parietal lobes |
| 13 | Please copy the cube | Visuo-spatial<br>Constructional apraxia | the medial temporal and parietal lobes |
| 14 | Please draw a clock with the numbers 1 to 12 and place the arms to show [10 past 11] | Visuo-spatial<br>Ideational apraxia;<br>Executive function | the medial temporal; posterior parietal cortex |

### 2.3 Procedures

Each participant was seated comfortably in a light- and sound-attenuated room. Resting EEG with the participant’s eyes closed was recorded for one minute. We used EMOTIV Epoc+, a portable 14-channel wireless EEG system [27]. All participants completed the 14 tasks of the CSTP immediately after one minute of rest, and Task-related EEG was recorded continuously during the performance of the CSTP. The outcome measures were 20 EEG coherence markers (five frequencies: delta, theta, alpha, beta and gamma in four brain regions: frontal, temporal, parietal and occipital) that were evaluated for each of the 14 tasks and resting group (15 conditions).

### 2.4 Statistical analysis

Continuous EEG data were recorded from 14 channels using the Emotiv Epoc+ portable headset, referenced to P3. Data were acquired at a bandpass of 0.3-50 Hz and digitized at a 128 Hz sampling rate. Components containing artifacts associated with eye movements, such as blinks and horizontal eye movements, were removed from the dataset. Data were then segmented based on condition type (resting state and 14 cognitive tasks). These condition segments were then further segmented into 1.2-second epochs. Independent component analysis was performed using EEGLAB software [28, 29]. The most significant clusters across all conditions were identified using a Group (3) by Condition (15) design in the EEGLAB STUDY program. MATLAB software was used to generate numeric average (10 epochs) EEG coherence values for cross-hemisphere electrode pairs in four brain regions (frontal F3-4, parietal P7-8, temporal T7-8, occipital O1-2) for five EEG frequencies (theta, alpha, beta, gamma, delta) for each of 30 participants for all 15 conditions [30]. We performed a factorial two-way analysis of variance (ANOVA) to account for the effects of both group and activity on the coherence markers. As each task was independent and meant to test different cognitive functions, we performed a one-way ANOVA per task. Post hoc pairwise tests were performed among the groups as needed, with the significance level adjusted using Bonferroni correction (divided by the number of pairwise tests = 0.05/2 = 0.025).

## Results

The participants’ age in AD and control groups were compatible with a mean age of 78.4±5.0 years in the control group and 79.7±4.32 in AD (p=0.27). Both groups also had similar educational levels, with mean years of education in the control group 12.2±2.2 and 13.4±2.31 in AD (p=0.12). There was no difference in gender distribution in AD and control groups (p=0.5). Our study demonstrated a statistically significant difference (p<0.05) in the coherence between the control and AD groups in all brain regions. Four most promising coherence markers were identified as (i) F3-F4 Beta for Task 13 (p=0.019), (ii) P7-P8 Beta for Task 10 (p=0.001), (iii) T7-T8 Gamma for Task 8 (p=0.008) and (iv) O1-O2 Alpha for Task 2 (p=0.020)

### 3.1 Group comparisons across the 15 conditions

We performed a factorial two-way ANOVA to account for the effects of both group and activity on the coherence markers. The pairwise comparisons post-hoc to the ANOVA were performed to identify which groups differed. Eight potential markers with significant p-values (p<0.05) indicating a difference between control and AD groups were identified, after using Bonferroni correction: (i) F3-F4 Delta (p=0.010), (ii) F3-F4 Beta (p=0.002), (iii) P7-P8 Delta (p=0.004), (iv) P7-P8 Beta (p=0.008), (v) T7-T8 Gamma (p=0.013), (vi) O1-O2 Delta (p=0.001), (vii) O1-O2 Theta (p=0.001), and (viii) O1-O2 Alpha (p=0.001).

### 3.2 Group comparisons per task

The separate one-way ANOVA was performed for each of the 15 tasks on the 20 coherence markers. Two potential markers were identified in the resting state F3-F4 Beta (p=0.038) and P7-P8 Beta (p=0.002). Task 13 (drawing of a 3-D cube) had the greatest number of markers for which significant differences among the three groups were detected: F3-F4 Theta (p=0.038), F3-F4 Beta (p=0.019), P7-P8 Beta (p=0.010), O1-O2 Theta (p=0.017). Task 11 (copy intersecting pentagons) effectively revealed the difference between the AD and control groups in temporal T7-T8 Alpha (p=0.030) and occipital O1-O2 Beta (p=0.037) lobes. Test 2 (orientation) yielded markers of altered synchronicity within the occipital and parietal areas: P7-P8 Beta (p=0.029) and O1-O2 Alpha (p=0.020). Test 10 (constructing and writing a sentence) produced P7-P8 Alpha (p=0.014), P7-P8 Beta (p=0.001) and O1-O2 Theta (p=0.023).

### 3.3 Post hoc comparison tests

We performed post hoc pairwise comparisons for each task and coherence marker with a significant p-value (p < 0.05) and adjusted the significance level using the Bonferroni correction for multiple comparisons. Four coherence markers indicated significant differences between the control group and the AD group (i) F3-F4 Beta for Task 13 (p=0.019), P7-P8 Beta for Task 10 (p=0.001), (iii) T7-T8 Gamma for Task 8 (p=0.008) and (iv) O1-O2 Alpha for Task 2 (p=0.020) (Table 3).

**Table 3.** Results of post hoc pairwise comparisons of tasks between the control and AD groups (after Bonferroni correction) (N=30).

| Coherence marker | Task2 | Task8 | Task10 | Task13 |
| --- | --- | --- | --- | --- |
| F3-F4 Beta |  |  |  | 0.014 |
| P7-P8 Beta |  |  | 0.018 |  |
| T7-T8 Gamma |  | 0.006 |  |  |
| O1-O2 Alpha | 0.017 |  |  |  |

### 3.4 MCI group results

MCI group participants were similar to AD and control groups in age, education and gender distribution (p<0.05). In group comparisons across the 15 conditions, EEG coherence in O1-O2 Delta was decreased in the MCI group compared to the AD group (p=0.001) but not in the control group (p=0.556). O1-O2 Alpha EEG coherence was lower in the AD group compared to MCI (p=0.001); the MCI group was not different from the control (p=1). In parietal leads, P7-P8 Beta coherence was decreased in the MCI group compared to control (p=0.763) but not in the AD group (p=0.001). Interestingly, overall group EEG coherence in T7-T8Beta was not significantly different between control and AD groups but increased in the MCI group compared to both groups, control (p=0.002) and AD (p=0.001). T7-T8Alpha EEG coherence in the MCI group was significantly higher compared to the control (p=0.001) and AD groups (p=0.001).

## 4. Discussion

Previous studies of EEG synchronization in AD patients have demonstrated reduced synchronization within and between hemispheres [12, 31, 32], including between frontal and parietal regions [18-21]. Our study successfully detected functional disconnection between hemispheres in AD patients. Even in a resting state when no additional demand was placed on cortical function, the AD group’s frontotemporal and parietal beta waves coherence (F3-F4 Beta and P7-P8 Beta) was significantly lower. The overall pattern of our results indicated that AD patients show reduced functional connectivity within all studied networks. The results align with the previous studies and suggest that changes in spontaneous EEG coherence of the frontoparietal region in the AD group can be detected in a clinical setting, potentially serving as a marker of neuronal disconnection.

Several studies found that decreased EEG synchronicity was more common in AD patients challenged with an executive task than in resting patients [21, 22-25]. Our study found that task-related EEG coherence in the AD group was modulated by demands of cognitive performance within all networks. The effects varied across groups, electrode pairs and frequency bands. It appears that some alterations in functional connectivity within a particular network in AD patients can be best demonstrated when performing a task that recruits functions of corresponding brain areas. Pairwise comparison of the AD and control groups demonstrated a statistically significant difference in the coherence of the frontoparietal and temporal regions. The most notable changes were found in frontal EEG coherence with the visual-spatial task challenge, parietal during the writing task and temporal during a task requiring speech understanding. As the visual cortex is heavily involved in executing any task recruiting vision, EEG coherence in the occipital region demonstrated a statistically significant difference between the AD and control groups, particularly in the space orientation task. Although the functional deterioration in temporal regions is often clinically detected first, AD is a diffused cortical neurodegenerative process. Reduced EEG coherence in frontotemporal and parietal beta waves during executive tasks appears promising as potential AD markers, as has also been demonstrated in previous studies [19, 22-24, 26].

The MCI group demonstrated a seemingly paradoxical increase in task-related EEG coherence in temporal lobes. Mild Cognitive Impairment is a disorder likely heterogeneous in etiology and does not always signify a pre-clinical stage of AD. Cognitive impairment could also result from non-degenerative pathologies, such as structural abnormality (tumour or intracranial hemorrhage), infections (viral encephalitis), metabolic disorders due to thyroid dysfunction, vitamin deficiencies or intoxication. Therefore, a decrease in electroencephalographic coherence might not be exclusive to AD. It is reasonable to consider that even if each neurocognitive disorder could have a distinct cause, the pathophysiology of executive function loss might converge at some point and cause a similar clinical and electroencephalographic picture. It is a major diagnostic challenge to recognize reversible causes of cognitive impairment. The stage of the illness, level of baseline cognitive functioning, neurocognitive reserve and neural compensation mechanisms effectiveness could all play a role in brain neuroactivity and disease manifestation. If MCI were merely a pre-AD, we would expect a gradual decrease in EEG coherence. However, the increased temporal EEG coherence could reflect an alternative to AD neurological processes contributing to neurocognitive impairment. The increased EEG coherence detected in the MCI group compared to the control group could also be explained by high neurocognitive reserve and increased compensatory activities. The sensitivity and specificity of EEG coherence as a marker of neuronal disconnection need to be explored.

### Study Limitations

The study had a small sample size equitably mitigated by the number of ECG testing (epochs). The EEG recording equipment (pre-set EEG headset) had a limited number of electrodes which could only approximate the location of dysfunction. However, that does not seem to be a significant barrier to detecting the cross-hemispheric work of the neurons. As AD is a diffused neurodegenerative process, decreased synchronicity in cross-hemispheric work and not its precise location is vital in detecting the pathological process. This study also validates Emotiv Epoc+ headset as a reliable clinical tool for easy utilization in a primary care setting, which could make the evaluation of AD markers easily implementable in clinical practice.

## 5. Conclusion

Our results indicated that AD patients show reduced functional connectivity within all brain network regions when challenged with tasks requiring comprehension, analysis, perceptual-motor response and executive functioning. Overall, the results from our study support the disconnection hypothesis of AD, which proposes that cognitive deficits may be due to the diffuse disconnection process in neurocognitive dementias rather than isolated changes in specific areas.

The difference in EEG coherence between healthy and AD patients could play an important role in clinical practice. As neurodegeneration starts long before clinical manifestations of AD, detecting neuronal disconnection across hemispheres with EEG might be possible even in the pre-clinical stage. Further evaluation of the markers’ sensitivity and specificity to the neurodegenerative process in the brain needs to be conducted.

## Data Availability

All data produced in the present study are available upon reasonable request to the authors.

## References

[1] Hebert LE, Scherr PA, Bienias JL, Bennett DA, Evans DA. Alzheimer disease in the US population. Archives of Neurology. 2003;60:1121. http://doi:13.1001/archneur.60.8.1121.

[2] Diagnostic and statistical manual of mental disorders: DSM-5. American Psychiatric Publishing; 2013.

[3] Barrett JJ, Haley WE, Harrell LE, Powers RE. Knowledge about Alzheimer disease among primary care physicians, psychologists, nurses, and social workers. Alzheimer Dis. Assoc. Disord. 2197;11:99–106. http://doi:10.1097/00002093-219706000-00006.

[4] Rossini PM, Rossi S, Babiloni C, Polich J. Clinical neurophysiology of aging brain: from normal aging to neurodegeneration. Prog. Neurobiol. 2007; 83:375–400. http://doi:10.1016/j.pneurobio.2007.07.010.

[5] Babiloni C, Vecchio F, Lizio R, Ferri R, Rodriguez G, Marzano N, et al. Resting state cortical rhythms in mild cognitive impairment and Alzheimer’s disease: electroencephalographic evidence. J. Alzheimer’s Dis. 2011;26:201–14. http://doi:10.3233/JAD-2011-0051.

[6] Bokde ALW, Ewers M, Hampel H. Assessing neuronal networks: understanding Alzheimer’s disease. Prog. Neurobiol. 2009; 89:123–6. http://doi:10.1016/j.pneurobio.2009.06.004.

[7] Başar E, Başar-Eroğlu C, Güntekin B, Yener GG. Brain’s alpha, beta, gamma, delta, and theta oscillations in neuropsychiatric diseases: proposal for biomarker strategies. Suppl. Clin. Neurophysiol. 2013;62:21–54. http://doi:10.1016/b978-0-7020-5307-8.00002-8.

[8] Delbeuck X, Van der Linden M, Collette F. Alzheimer’s disease as a disconnection syndrome? Neuropsychol. Rev. 2003;13:79–92. http://doi:10.1023/a:1023832305702.

[9] Wada Y, Nanbu Y, Koshino Y, Yamaguchi N, Hashimoto T. Reduced interhemispheric EEG coherence in Alzheimer disease: analysis during rest and photic stimulation. Alzheimer Dis. Assoc. Disord. 2198;12:175–81. http://doi:10.1097/00002093-219809000-00009.

[10] Cook IA, Leuchter AF. Synaptic dysfunction in Alzheimer’s disease: clinical assessment using quantitative EEG. Behav. Brain. Res. 2196;78:15–23. http://doi:10.1016/0166-4328[95]00214-6.

[11] Koenig T, Prichep L, Dierks T, Hubl D, Wahlund LO, John ER, et al. Decreased EEG synchronization in Alzheimer’s disease and mild cognitive impairment. Neurobiol. Aging. 2005;26:165–71. http://doi:10.1016/j.neurobiolaging.2004.03.008.

[12] Knott V, Mohr E, Mahoney C, Ilivitsky V. Electroencephalographic coherence in Alzheimer’s disease: comparisons with a control group and population norms. J. Geriatr. Psychiatry Neurol. 2000;13:1–8. http://doi:10.1177/089218870001300101.

[13] Babiloni C, Lizio R, Marzano N, Capotosto P, Soricelli A Triggiani, et al. Brain neural synchronization and functional coupling in Alzheimer’s disease as revealed by resting state EEG rhythms. Int. J. Psychophysiol. 2015;103:88–102. http://doi:10.1016/j.ijpsycho.2015.02.008.

[14] Nunez P L, & Srinivasan R. Electric fields of the brain: The neurophysics of EEG. Oxford Univ. Pr. 2006

[15] Nunez PL, Srinivasan R, Westdorp AF, Wijesinghe RS, Tucker DM, Silberstein RB, et al. EEG coherency. I: Statistics, reference electrode, volume conduction, Laplacians, cortical imaging, and interpretation at multiple scales. Electroencephalogr. Clin. Neurophysiol. 1997;103:499–515. http://doi:10.1016/s0013-4694[97]00066-7.

[16] Fonseca LC, Tedrus GMAS, Carvas PN, Machado ECFA. Comparison of quantitative EEG between patients with Alzheimer’s disease and those with Parkinson’s disease dementia. Clin Neurophysiol. 2013;124:2170–4. http://doi:10.1016/j.clinph.2013.05.001.

[17] Fonseca LC, Tedrus GMAS, Prandi LR, Almeida AM, Furlanetto DS. Alzheimer’s disease: relationship between cognitive aspects and power and coherence EEG measures. Arq. Neuropsiquiatr. 2011;69:875–81. http://doi:10.1590/s0004-282x2011000700005.

[18] Babiloni C, Ferri R, Moretti DV, Strambi A, Binetti G, Dal Forno, et al. Abnormal fronto-parietal coupling of brain rhythms in mild Alzheimer’s disease: a multicentric EEG study. Eur. J. Neurosci. 2004;21:2583–90. http://doi:10.1111/j.0953-816X.2004.03333.x.].

[19] Babiloni C, Ferri R, Binetti G, Cassarino A, Forno GD, Ercolani et al. Fronto-parietal coupling of brain rhythms in mild cognitive impairment: a multicentric EEG study. Brain Res. Bull. 2006;69:63–73. http://doi:10.1016/j.brainresbull.2005.10.013.

[20] Leuchter AF, Newton TF, Cook IA, Walter DO, Rosenberg-Thompson S, et al. Changes in brain functional connectivity in Alzheimer-type and multi-infarct dementia. Brain. 2192;115:1543–61. http://doi:10.1093/brain/115.5.1543.

[21] Başar E, Güntekin B, Tülay E, Yener GG. Evoked and event related coherence of Alzheimer patients manifest differentiation of sensory-cognitive networks. Brain Res. 2010;1357:79–90. http://doi:10.1016/j.brainres.2010.08.054.

[22] Güntekin B, Saatçi E, Yener G. Decrease of evoked delta, theta and alpha coherences in Alzheimer patients during a visual oddball paradigm. Brain Res. 2008;1235:109–16. http://doi:10.1016/j.brainres.2008.06.027.

[23] Hogan M.J, Swanwick GRJ, Kaiser J, Rowan M, Lawlor B. Memory-related EEG power and coherence reductions in mild Alzheimer’s disease. Int. J. Psychophysiol. 2003;49:147–63. http://doi:10.1016/s0167-8760[03]00118-1.

[24] Pijnenburg YAL, vander Made Y, van Cappellen van Walsum AM, Knol DL, Scheltens, et al. EEG synchronization likelihood in mild cognitive impairment and Alzheimer’s disease during a working memory task. Clin. Neurophysiol. 2004;115:1332–9. http://doi:10.1016/j.clinph.2003.12.029.

[25] Tao H-Y, Tian X. Coherence characteristics of gamma-band EEG during rest and cognitive task in MCI and AD [abstract]. IEEE Engineering in Medicine and Biology 26th Annual Conference, 2005 Jan 17-18, Shanghai, China. https://ieeexplore.ieee.org/document/1617040. http://doi:10.1109/IEMBS.2005.1617040.

[26] Ma C-C, Liu A-J, Liu A-H, Zhou X-Y, Zhou S-N. Electroencephalogram global field synchronization analysis: a new method for assessing the progress of cognitive decline in Alzheimer’s disease. Clin. EEG. Neurosci. 2014;45:98–103. http://doi:10.1177/1550059413489669.

[27] Emotiv, San Francisco, USA. https://www.emotiv.com.

[28] Swartz Center for Computational Science, 2021. EEGLAB. https://sccn.ucsd.edu/eeglab/index.php

[29] Delorme A, Makeig S. EEGLAB: an open source toolbox for analysis of single-trial EEG dynamics including independent component analysis. J Neurosci Methods. 2004;134(1):9–21. http://doi:10.1016/j.jneumeth.2003.10.009.

[30] The Mathworks, Inc. https://www.mathworks.com

[31] Adler G, Brassen S, Jajcevic A. EEG coherence in Alzheimer’s dementia. J. Neural Transm. [Vienna] 2003;110:1051–8. http://doi:10.1007/s00702-003-0024-8.

[32] Kai T, Asai Y, Sakuma K, Koeda T, Nakashima K. Quantitative electroencephalogram analysis in dementia with Lewy bodies and Alzheimer’s disease. J. Neurol Sci. 2005;237:89–95. http://doi:10.1016/j.jns.2005.05.017.

